# Revisiting the plague epidemic in Gévaudan, 1720-1722: the key roles of multiple zoonotic introductions and human-to-human transmission

**DOI:** 10.1101/2025.07.18.25331563

**Authors:** Thibaut Jombart, Anne Cori, Lilith K Whittles, Henry Mouysset

**Affiliations:** MRC Centre for Global Infectious Disease Analysis, School of Public Health, Imperial College London, United Kingdom; Retired

## Abstract

We revisited the spread of the plague epidemic which decimated the province of Gévaudan, France, in 1720-1722, and was assumed to have its origin in the great plague epidemic of Marseille, 1720. Using historical records of bubonic plague deaths in the first affected village of Corréjac, we showed that the commonly admitted origin of the epidemic, thought to have been seeded by an escaped convict from Marseille, is highly implausible. We developed a statistical model to estimate the respective intensities of human-to-human and rodent-to-human transmission, and showed that both pathways were needed to sustain the epidemic, with person-to-person transmission accounting roughly for two thirds of all cases. These results bring support to the theory that human ectoparasites such as fleas and lice may play an important role in bubonic plague epidemics through the mediation of human-to-human transmission.

## INTRODUCTION

A fundamentally applied science, infectious disease epidemiology mostly focuses on recent epidemics as well as on future potential public health threats. However, the study of historical outbreaks also has its own value, not only to better understand how these events have shaped societies [1–4], but also because the dynamics of major historical epidemics may yet inform us on the spreading potential and impact of current and future pathogens [4– 9]. This is especially true of the bacterium *Yersinia pestis*, the causative agent of plague, which has been responsible for the most terrible epidemics in recorded history [3,5,10,11], and remains to this day a pathogen of substantial concern, despite the advent of antibiotic treatments [11–14].

Plague manifests in three clinical forms - bubonic, septicemic, and pneumonic - distinguished by the primary site of infection, mode of transmission, and severity [15]. Bubonic plague, the most common form, is typically introduced into human populations from zoonotic reservoirs (rodent populations [16]) through the bites of infected fleas [17]. Once established in humans, the disease can spread further either indirectly, through human ectoparasites such as body lice or human fleas, or, if the infection progresses to pneumonic plague, directly between individuals via respiratory droplets [15]. The relative importance of zoonotic and person-to-person transmission routes in historical epidemics remains debated. Traditionally, bubonic outbreaks were thought to have been driven primarily by rodent-to-human transmission mediated by rat fleas [18]. However, modeling studies, including analyses of the Eyam outbreak of 1665-66 [19], European outbreaks of the second pandemic [20], and the Glasgow outbreak of 1900 [21], suggest that human ectoparasites may have played a central role in sustaining human-to-human bubonic plague transmission.

The plague epidemic in Marseille, France, in 1720-1722 was one of the last great European epidemics. Thought to have been introduced by the ship ‘Grand Saint Antoine’ sailing from Cyprus on the 25th of May 1720 [22,23], it had devastating effects on the population: out of about 90,000 inhabitants, 10,000 fled the city, and about half of the remaining population died [24]. Importantly, the epidemic did not stop in Marseille, but is thought to have spread despite blockades put in place to about 240 cities in Provence and Languedoc during the fall of 1720 [22,23], resulting in a staggering overall death toll of 120 000 deaths out of an estimated population of 400 000 inhabitants [22], with larger cities often more severely affected [24]. In Gévaudan alone, a province of about 90,000 inhabitants which was key to the wool industry and trade, the plague epidemic virtually put wool-related activities and commerce to a stop, and decimated entire towns with death tolls exceeding 60% of the population in some cities [25].

While it is generally thought that the plague spread via trade routes [24], the origin of the plague epidemic in Gévaudan remains unclear. The commonly accepted scenario is that a convict assigned to burying bodies in Marseille escaped the city carrying a flea-infested bundle of wool, which he would have then sold at a wool market some 300 km away in Saint-Laurent-d’Olt, on the 23^rd^ November 1720 [25]. His buyer, Jean Quintin (JQ), later known as the index case of the plague epidemic in Gévaudan, started showing symptoms of bubonic plague within the next 24 hours. He came back to his home in the town of Corréjac, where he infected his son, seeding an epidemic which rapidly took over the village, nearby towns, and eventually, the entire province [25]. Several aspects of this scenario are, however, debatable. The idea that a convict could have escaped Marseille and travelled such a distance on foot while avoiding blockades, and remaining uninfected the entire trip, is in direct contradiction with the observation that most convicts who were put on burial duty showed symptoms within a few days of work and subsequently died (mortality 94%, *n* = 217 [24]). And beyond this questionable introduction, the modes of transmission (rodent-to-human *versus* human-to-human) and general dynamics which enabled *Y. pestis* to spread to the whole province remain unknown.

Here, we analyse historical records of bubonic plague deaths to revisit the origin and spread of the plague epidemic in Gévaudan. First, we test the plausibility of the ‘convict’ introduction scenario, estimating the daily rate of infection from the wool bundle and the overall probability of the initial transmission events. Second, we introduce a disease transmission model which, combined with historical data on deaths from bubonic plague in Corréjac, is used to estimate the respective roles of person-to-person transmission and zoonotic introductions in the spread of plague in Gévaudan.

## MATERIAL AND METHODS

### Data

Data on the date of deaths of bubonic plague cases were collected from historical records in Corréjac from November 1720 to August 1721. Deaths were considered to be plague-related whenever records referred to the deceased as having exhibited buboes, typically alongside high fever, and extreme weakness. Historical records of the plague epidemic in Corréjac mention no plague survivor, and while such isolated cases have been later documented in the Gévaudan epidemic, they remained extremely rare [25]. We therefore assumed plague deaths essentially capture all cases in Corréjac.

A few early cases assigned to the nearby town of La Canourgue, belonging to the same family as index case JQ and for which a clear epidemiological link to the index case was identified, were also included. The final dataset consisted of 52 cases, including 43 cases from Corréjac, and 9 cases from La Canourgue.

### Delay distributions

Analysis of the timing of epidemiological events required information on the incubation period (time from infection to onset of symptoms) and the duration of illness (time from the onset of symptoms to death) distributions. The incubation period distribution was characterized as a discretized lognormal distribution with mean 3.5 days and standard deviation (SD) 1.5 days (Figure S1) to match historical data reporting delays of 2 to 6 days between infection and the onset of symptoms of bubonic plague [26–32]. We denote the corresponding daily probability mass function (PMF) as. Similarly, the duration of illness distribution was characterized as a discretized lognormal distribution with mean 3 days and SD 1.4 days (Figure S2) to match data from the literature reporting that most bubonic plague deaths in the absence of treatment occurred within 3 to 5 days of the symptoms onset [21,26,31,33]. We denote the corresponding PMF as.

Our transmission model also required characterization of the generation time distribution (PMF:), *i.e.* the time interval between a primary infection and onward transmission. We derived this distribution from the incubation period and the duration of illness, assuming that they are independent and that transmission takes place with uniform probability during the symptomatic period. This was achieved by numerically approximating the distribution of infectious periods *i.e.* the convolution of the incubation and symptomatic periods, by drawing 10 million samples from the distributions described above, then drawing transmission times uniformly between the date of infection and death. The full approach is described in Supplementary Text S1, and the resulting distribution shown in Figure S3.

### Likelihood of initial transmissions

The seeding of the plague epidemic in Gévaudan has two aspects, which were evaluated separately: first, the infection of JQ by a convict travelling from Marseille via an infected wool bundle, and second, the infection of JQ’s son by his father.

We first assess the likelihood of the convict traveling with an infected wool bundle which subsequently infected JQ within 24 hours of his purchase. Assuming the daily rate of infection from the wool bundle is constant over time and *D* is the duration of the convict’s travel from Marseille (during which no transmission occurred), the likelihood of the initial transmission is given by the geometric distribution:

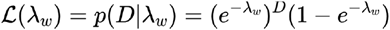

where the first term is the probability of no transmission during *D* days, and the second is the probability of transmission on day *D* + 1. As *D* is not known, we have considered 3 different scenarios, for an assumed distance of around 300 km: a short travel (7 days, > 40km per day), an average one (12 days, ≃25km per day), and a longer one (20 days, ≃15 km per day). The likelihood profile was derived for a regular grid of 1,000 values of, ranging from 0 to 1.2 per day for each scenario.

Second, we examined the hypothesis of a direct transmission from JQ (sick on the 23^rd^, dead on 27^th^ November 1720) to his son (dead on the 18^th^ December 1720, date of symptom onset unknown), by evaluating the plausibility of the delay from infection to death of the son. Because transmission could have occurred between the 24^th^ and 26^th^ November 1720, we considered 3 different possible delays from infection to death, from 24 to 26 days. Expected delays from infection to deaths were obtained by adding simulated incubation times (Figure S1) to simulated duration of illness (Figure S2). This operation was performed 10,000,000 times to obtain an empirical distribution to which observed delays were compared, and for which associated *p*-values were computed as the corresponding quantiles.

As *Y. pestis* has been shown to survive on infected clothes or bed sheets for up to 5 days [34], we also performed a sensitivity analysis assuming contamination could have taken place up to 5 days after JQ’s death, calculating *p*-values associated to delays from infection to death of 19-21 days.

### Transmission model

We used a branching process approach to characterize the disease transmission, as these models are particularly effective at estimating transmissibility from time series of case incidence [35–38]. A Hawkes process [39] was used to account for person-to-person transmission as well as zoonotic introductions, using augmented data [40–42] to integrate over all possible (unobserved) dates of infections, given the (observed) dates of death. We denote the total number of cases in the outbreak as *n*, and its duration in days as *T*, calculated as the time interval between the first and last case of the outbreak.

Our approach is embedded in a Bayesian framework where the posterior distribution is defined as:

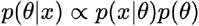

where *θ* refers to the parameters, *p*(*x*|θ) is the data, is the likelihood function and *p*(θ) the joint priors. Our model’s likelihood is defined as (see details in Text S2):

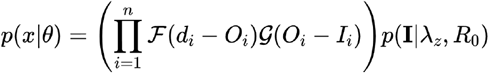

where *d*_*i*_ is the date of death of case *i, O*_*i*_ and *I*_*i*_ are augmented data respectively corresponding to the date of onset and infection, λ_*z*_ is the daily rate of zoonotic introduction (rodent-to-human transmission), and *R*_0_ is the basic reproduction number, *i.e.* the average number of secondary infections caused by each case in a fully susceptible population (human-to-human transmission). Bold font is used to indicate vectors, with *I*={*I*_*i*_} ∀ *i*=1,…,*n*.

The first term between parentheses indicates the probability of the duration of illness and incubation times for all cases. The second term can be reformulated as the probability of the case incidence given the disease’s transmissibility and zoonotic introductions:

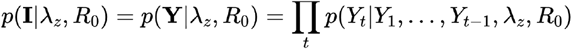

where *Y* = {*Y*_*t*_} ∀ *t* = 1,…,*T* and where *Y*_*t*_ is the case incidence (by date of infection) at time *t*:

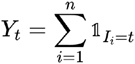

where 𝟙 is the indicator function. The probability of the case incidence *Y*_*t*_ is defined by the Hawkes branching process [35,39] with :

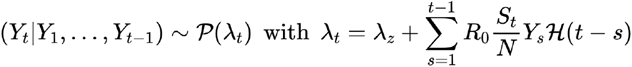

where *P*(·) is the Poisson distribution, is the rate of infection at time *t, S*_*t*_ is the number of remaining susceptible individuals in the population at time *t*, and *N* is the total population size of Corréjac, estimated to 111 inhabitants [25]. This modification of usual branching process models [35–37] accounts for the depletion of susceptible individuals in the population, an important factor here since 47% of the village’s population died during the epidemic.

We assume independent priors for λ_*z*_ and *R*_0_. We used a prior for *R*_0_ derived from a 1900 bubonic plague epidemic in Glasgow [21] a lognormal distribution with mean 1.6 and SD 1.3 (Figure S4). In the absence of information on the rate of zoonotic introductions, we used a flat, uninformative uniform prior ranging from 0 to 10.

### Estimation process

The Metropolis algorithm [43] was used to derive samples from the posterior distribution (see details in Supplementary Text S2). The model and estimation process were implemented in the R software [44]. A total of 12 independent chains were run in parallel for 10,000 iterations each, using random initial states. Convergence was assessed visually (Figure S5), and effective sample size calculations were used to confirm that a thinning of 1 in 10 would yield little to no pseudoreplication. A burnin of 100 steps was used (Figure S5). The final sample size for all posterior estimates was therefore 4,752. Parameters were first estimated using cases from Corréjac only, and then also including epidemiologically related cases from La Canourgue as a sensitivity study. To compare the respective impact of human-to-human and rodent-to-human transmission, we translated estimated values of *R*_0_ into an average daily rate of human-to-human infection in a fully susceptible population as:

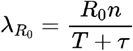

where is the mean generation time, derived from our simulated distribution (Text S1).

### Data Availability

The data and scripts implementing the model and all analyses presented in this paper are publicly available from the github repository: https://github.com/thibautjombart/gevauplague

## RESULTS

The general timeline and epidemic curve of the bubonic plague epidemic in Corréjac are shown in Figure 1. Estimations of the daily rate of infection from the wool bundle show that low rates were most likely for all scenarios (Figure 2), with maximum likelihood estimates (MLE) of λ_*w*_ ranging from 0.0949 per day (for the short travel duration scenario: 10 days) to 0.0649 (15 days), and 0.0492 (20 days). The corresponding probabilities of each scenario, based on the respective MLEs, were low, ranging from 1.8% for a trip of 20 days to 3.5% for a trip of 10 days. The hypothesis of a direct transmission from JQ to his son also proved highly improbable (Figure 3). Our simulations showed that the delay from all possible dates of infection of JQ’s son to his death was highly unlikely (*p*-values ranging from 1.95×10^−5^ to 5.80×10^−6^), even when considering the possibility of indirect transmission via infected fabric up to 5 days after JQ’s death (*p*-values ranging from 3.93×10^−4^ to 1.10×10^−5^). Taken together, these results suggest that the commonly accepted origin of the plague epidemic in Gévaudan is highly implausible.

**Figure 1.**
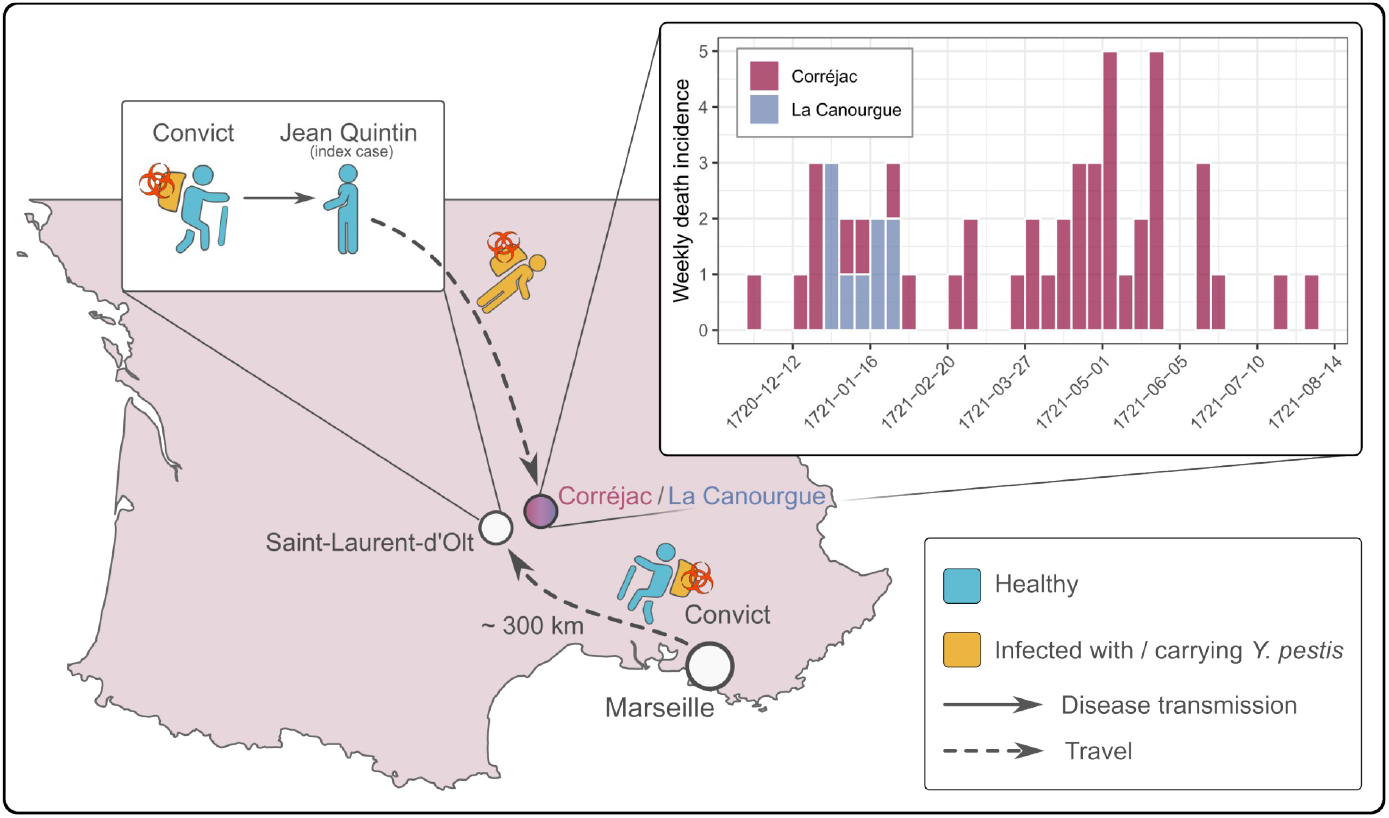
Timeline of the bubonic plague in Corréjac. This diagram summarises the hypothetical scenario of the beginning of the bubonic plague in Corréjac and La Canourgue. A convict on forced burial duty in Marseille would have escaped with a *Y. pestis-*infected wool bundle, walked some 300 km to Saint-Laurent-d’Olt without getting infected, and would have sold the wool bundle to Jean Quintin, the index case of the epidemic, who showed symptoms on the day of purchase and died 3 days later, after seeding the epidemic. The epidemic curve on the top-right of the figure shows weekly incidence by date of death, colored by the town of the deceased. Cases from La Canourgue had a clear epidemiological link with the index case (same family).

**Figure 2.**
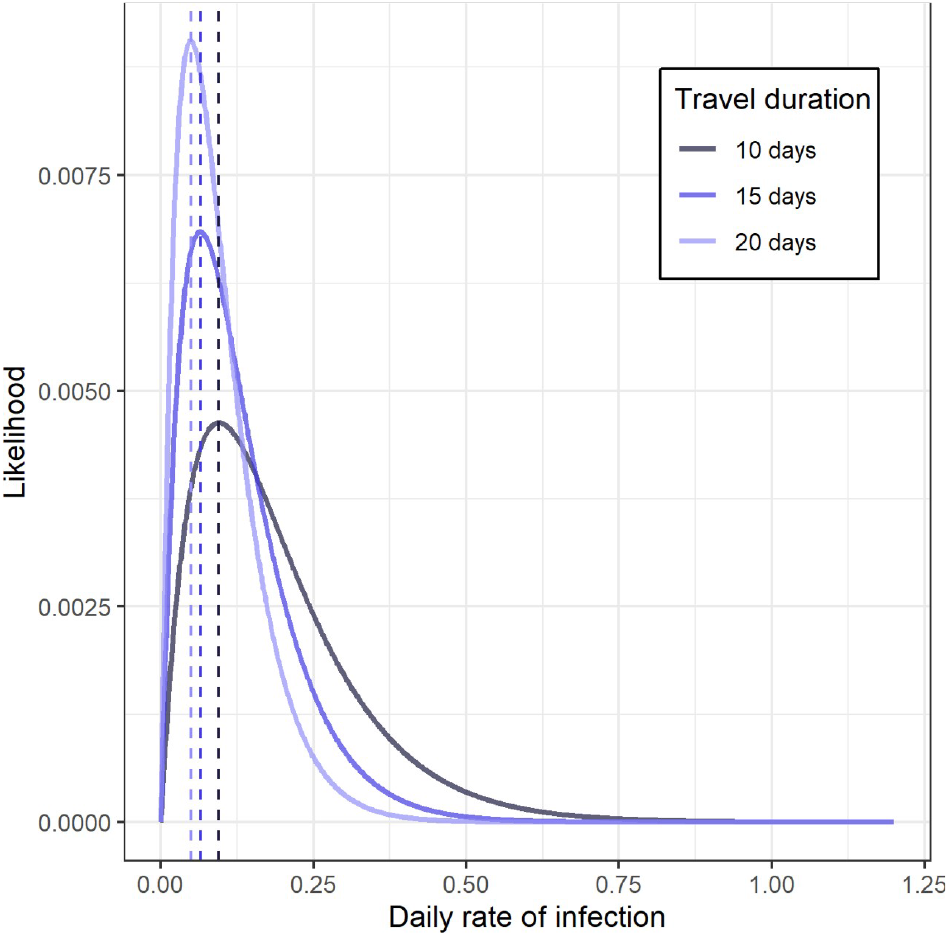
Estimation of the daily rate of infection from the wool bundle. The different curves plot the likelihood profiles of different rates of daily infection from the wool bundle according to different lengths of travel from Marseille to Saint-Laurent d’Olt. Vertical lines indicate the maximum likelihood estimates of 0.0949 (10 days), 0.0649 (15 days), and 0.0492 (20 days).

**Figure 3.**
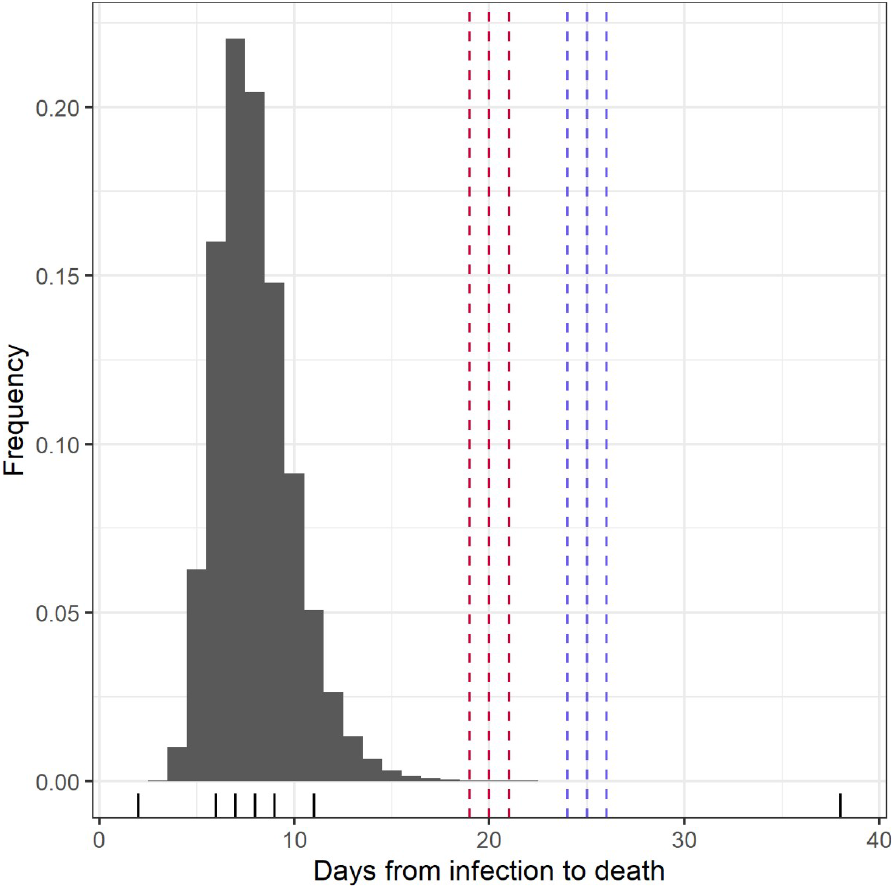
Estimation of plausible delays from infection to death of the first secondary case. The histogram shows the frequency distribution of delays from infection to death based on 10 millions simulations. Tick marks indicate the deciles of the distribution. The dashed vertical lines indicate the possible delays from infection to death of JQ’s son, assuming direct transmission (blue, 24-26 days), or from infected bed sheets or clothes after JQ’s death (red, 19-21 days).

Results from our transmission model provide further insights into the initial spread of the bubonic plague in Corréjac (Figure 4). The generation time distribution was estimated with a mean of 4.89 days and standard deviation 1.98 days (Text S1, Figure S3). The estimated values of *R*_0_ for Corréjac were substantially lower than priors derived from the literature, with an average of 0.95 (95% credible intervals, 95%_CrI_: [0.67; 1.28]), and only 36.2% of posterior values of *R*_0_ exceeding 1 (Figure 4A). This result implies that the spread of the disease in Gévaudan is unlikely to have taken place due to person-to-person transmission alone. Accordingly, our model estimated a low, but non-negligible daily rate zoonotic introduction averaging 0.08 (95%_CrI_ = [0.04; 0.13]), approximately corresponding to 1 introduction from the zoonotic reservoir every 13 days (95%_CrI_: 8-25 days). When calculating the respective contributions of the different sources of infection, we found human-to-human transmission accounted on average for 68% of all transmissions (95%_CrI_ = [51%; 83%]), while conversely rodent-to-human transmissions represented 32% of all transmissions (95%_CrI_ = [17%; 49%]).

**Figure 4.**
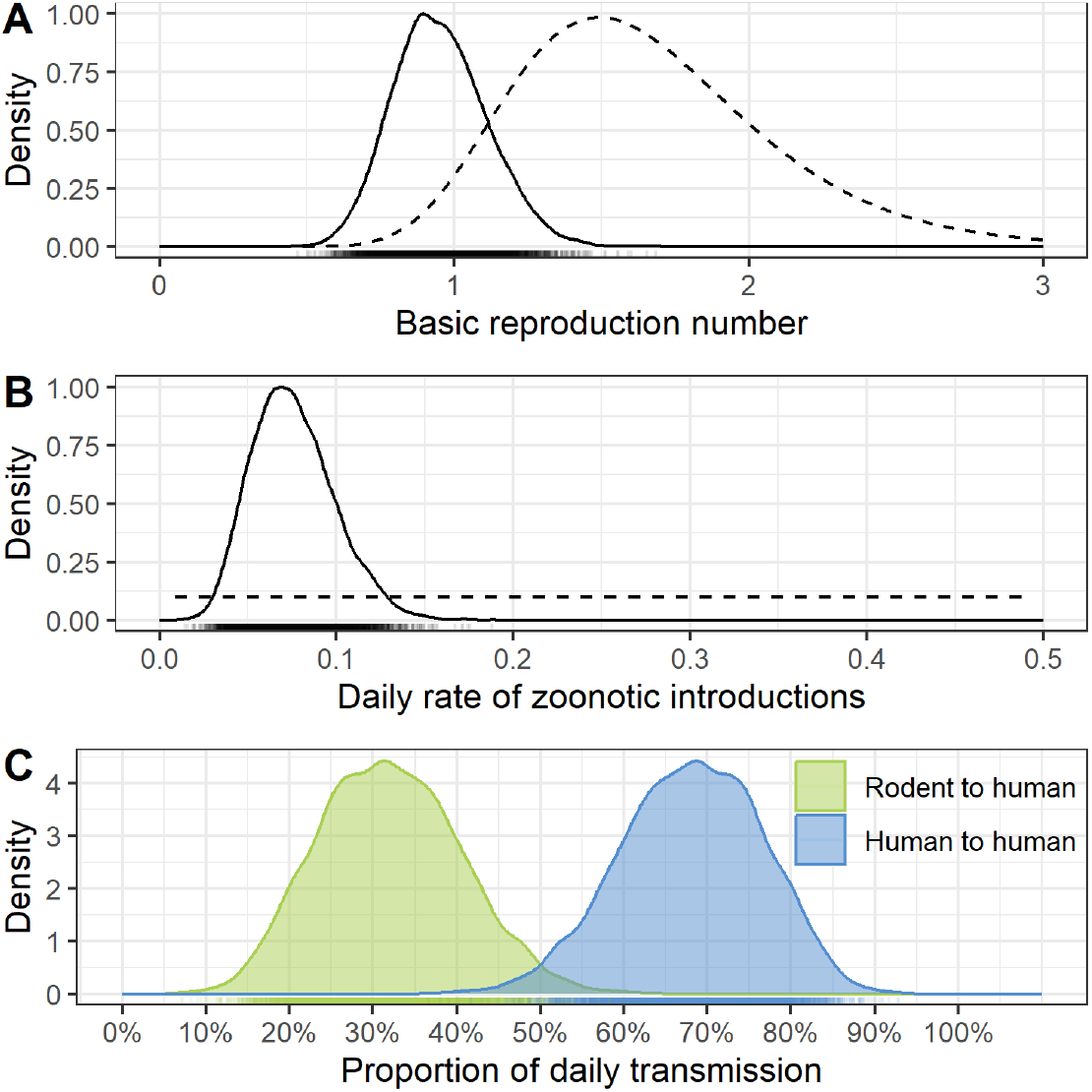
Estimation of transmissibility by source of infection. Plain curves represent the kernel density estimation of the posterior samples of the model parameters merged from 12 independent chains. Each chain was run for 10,000 iterations, with a burnin of 100 iterations and a thinning of 1/25, resulting in a total of 4,752 posterior samples. Dashed lines represent priors. **A)** Basic reproduction number *R*_0_ for human-to-human transmission **B)** Daily rate of zoonotic introductions. **C)** Proportion of the daily force of infection from the two respective sources. Semi-transparent tick marks indicate individual samples.

These results remained essentially identical when including epidemiologically related cases from La Canourgue, with an average *R*_0_ of 0.99 (95%_CrI_: [0.69; 1.32]) and a mean of 0.08 (95%_CrI_ = [0.04; 0.14]). With these cases included, the relative contributions of the sources of infection remained similar, with 70% of human-to-human transmissions (95%_CrI_ = [53%; 86%]).

## DISCUSSION

We combined modern statistical approaches with historical death records to revisit the spread of the bubonic plague in Gévaudan in 1720. Our results suggest that the generally accepted scenario of a convict escaping Marseille with an infected wool bundle, and seeding a new epidemic some 300km away in Gévaudan, is unlikely, even if we cannot formally rule out that the convict could have been a plague survivor, and therefore immune to the infection. Similarly, the long delay between the death of JQ and his son is highly incompatible with a direct transmission event, and rather suggests a rodent-to-human infection. While the traditional origin story of the plague epidemic in Gevaudan is unlikely to be plausible, the true beginnings of the outbreak remain an open question.

Further results from our modelling study show that the epidemic could only be explained by a combination of rodent-to-human and human-to-human transmission. Indeed, person-to-person transmission alone could not have sustained such a long epidemic due to a likely subcritical basic reproduction number (*R*_0_ = 0.95, 95%_CrI_: [0.67; 1.28]), and recurrent zoonotic introductions, albeit at a low rate (roughly 1 introduction every 1-3 weeks), were instrumental in spreading the disease further.

Our results are congruent with observations made on the Eyam bubonic plague of 1665-1666, where both rodent-to-human and human-to-human transmission routes were instrumental in sustaining the epidemic, estimated to have respectively accounted for 25% and 75% of transmissions [19], broadly similar to the 32% and 68% of transmissions in our study. It should be emphasized that, as in the Eyam outbreak study, our definition of ‘human-to-human’ transmission includes ectoparasite-mediated transmission [20,45], so that our results bring support to the theory that human lice [46,47] or fleas [48,49] may play a key role in bubonic plague dynamics.

A number of limitations need to be taken into account when considering the results of this study. First, our model assumes that there were no cases unreported. Historical death records for Corréjac and La Canourgue were generally very comprehensive, so that we are confident that the vast majority of bubonic plague deaths were recorded in the early stages of the Gévaudan epidemic. We cannot exclude that some cases went unnoticed, either because some deaths might not have been attributed to plague, or because some patients may have survived. We expect that our results should remain essentially unaffected as long as unreported cases were marginal.

Another potential issue is that our model does not account for potential imports of cases: secondary cases are assumed to only happen through local human-to-human or rodent-to-human transmission. In theory, imported cases could bias our estimates of transmissibility, if they represented a substantial fraction of cases. We believe the likelihood that there were multiple introductions is, however, very low. First, the only known potential source at the beginning of the outbreak was Marseille, and our results show that a single introduction from this source was unlikely in the first place. Second, roadblocks and containment of entire villages once they were known to be affected by plague would have drastically limited travel in the later stages of the outbreak [25], so that reintroductions most likely played a very limited role in the overall dynamics of the plague epidemic in Gévaudan.

Lastly, we only focussed on the early stages of the epidemic in the initial foci of Corréjac, mostly due to insufficient data in historical records for a broader study. While our results may be a good reflection of the beginning of the outbreak and its initial spread throughout Gévaudan, it is possible that the dynamics of plague transmission may have changed in later stages and different settings. For instance, in cities under lockdown but with poor sanitation and large murine populations, it is possible that the rodent-to-human transmissions would have played a more important, or even dominant role.

## CONCLUSION

Our study suggests that the plague epidemic in Gévaudan in 1720-1722 was driven by a mixture of person-to-person and zoonotic transmissions, bringing support to theories that human ecto-parasites may play a key role in bubonic plague outbreaks. Our results are entirely reproducible and the model we have developed can easily be adapted to other outbreak and disease contexts.

## ACKNOWLEDGEMENTS

AC, LKW, and TJ acknowledge funding from the MRC Centre for Global Infectious Disease Analysis (reference MR/X020258/1), funded by the UK Medical Research Council (MRC). This UK funded award is carried out in the frame of the Global Health EDCTP3 Joint Undertaking. LKW acknowledges funding from the Wellcome Trust [grant number 218669/Z/19/Z].

