## Supplemental Material for "Revisiting the plague epidemic in Gévaudan, 1720-1722: the key roles of multiple zoonotic introductions and human-to-human transmission"

**Text S1: Generation time distribution of bubonic plague**

The generation time distribution *H* was derived numerically using the following procedure:

1. Sample a large number *k* of incubation periods *X* using a continuous, lognormal distribution with mean 3.5 and SD 1.5 days
2. Sample a large number *k* of infectious period durations *Y* using a continuous, lognormal distribution with mean 3 and SD 1.4 days
3. Sample *k* delays from onset to infections *Z*, uniformly distributed between 0 and *Y*
4. Derive the empirical distribution of $H = f(X + Z)$ where *f* refers to the flooring function

This procedure was carried out with $k = 10^7$ draws. Further increasing the number of draws did not change substantially the quartiles, average, or median of the resulting distribution.

**Text S2: a Hawkes process model for bubonic plague transmission**

*Notations*

We use the following notations for the data, parameters, and distributions:

- $i = 1, \ldots, n$: index of individuals
- $t = 1, \ldots, T$: time index
- $d_i$: date of death of case $i$ (data) ; we note the vector $\mathbf{d} = \{d_i\} \ \forall \ i = 1, \ldots, n$
- $O_i$: date of symptom onset of case $i$ (augmented data) ; we note the vector $\mathbf{O} = \{O_i\} \ \forall \ i = 1, \ldots, n$
- $I_i$: date of infection of case $i$ (augmented data) ; we note the vector $\mathbf{I} = \{I_i\} \ \forall \ i = 1, \ldots, n$
- 
- $Y_t$: incidence of new infections at time $t$ (augmented data), derived from $I_i$ as

  $$Y_t = \sum_{i=1}^{n} \mathbb{1}_{I_i = t}$$

  We note the vector $\mathbf{Y} = \{Y_t\} \ \forall \ t = 1, \ldots, T$
- $S_t$: the number of susceptible (*i.e.* non-infected) individuals at time $t$, calculated as $S_t = N - \sum_{s=0}^{t} Y_s$
- $N$: the total number of individuals (infected and non-infected) in the area considered, assumed constant over the span of the epidemic; includes deaths from the disease
- $\lambda_z$: the rate of zoonotic introduction, assumed constant over time (parameter)
- $R_0$: the basic reproduction number (parameter)
- $\mathcal{F}$: probability mass function (pmf) of the infectious/symptomatic period distribution
- $\mathcal{G}$: pmf of the incubation period distribution
- $\mathcal{H}$: pmf of the generation time distribution, obtained as described in text S1

*Posterior distribution and likelihood*

Our approach is embedded in a Bayesian framework where:

$$p(\theta|x) \propto p(x|\theta)p(\theta)$$

where $\theta$ refers to the parameters, $x$ is the data, $p(x|\theta)$ is the likelihood function and $p(\theta)$ the joint priors.

Our model's likelihood is defined as:

$$
\begin{aligned}
p(x|\theta) &= p(\mathbf{d}, \mathbf{O}, \mathbf{I}|\lambda_z, R_0) \\
&= p(\mathbf{d}|\mathbf{O})p(\mathbf{O}|\mathbf{I})p(\mathbf{I}|\lambda_z, R_0) \\
&= \left(\prod_i p(d_i|O_i) \prod_i p(O_i|I_i)\right) p(\mathbf{I}|\lambda_z, R_0) \\
&= \left(\prod_i \mathcal{F}(d_i - O_i)\mathcal{G}(O_i - I_i)\right) p(\mathbf{I}|\lambda_z, R_0)
\end{aligned}
$$

The calculation of $p(\mathbf{I}|\lambda_z, R_0)$ is defined by the Hawkes process for the incidence **Y**:

$$
\begin{aligned}
p(\mathbf{I}|\lambda_z, R_0) &= p(\mathbf{Y}|\lambda_z, R_0) \\
&= \prod_t p(Y_t|Y_1, \dots, Y_{t-1}, \lambda_z, R_0)
\end{aligned}
$$

The incidence $Y_t$ is governed by:

$$
Y_t|Y_1, \dots, Y_{t-1} \sim \mathcal{P}(\lambda_t)
$$

where $\mathcal{P}(.)$ is the Poisson distribution, and with:

$$
\lambda_t = \lambda_z + \sum_{s=1}^{t-1} R_0 \frac{S_t}{N} Y_s \mathcal{H}(t - s)
$$

Note that we assume that $p(Y_1)$ is constant. Finally, we assume independent priors for $\lambda_z$ and $R_0$ (see section on Priors below) such that:

$$
p(\theta) = p(\lambda_z, R_0) = p(\lambda_z)p(R_0)
$$

*Priors*

We used a prior for $R_0$ derived from a 1900 bubonic plague epidemic in Glasgow [1] as a lognormal distribution with mean 1.6 and SD 1.3 (Figure S4). In the absence of information on the daily rate of zoonotic introductions, we used a flat, uninformative prior as a uniform distribution ranging from 0 to 10.

*Estimation process*

We sample from the posterior distribution using the Metropolis algorithm with augmented data using the following procedure:

1. Propose new augmented data $\mathbf{O}^*$ using $d_i - O_i^* \sim \mathcal{F}$ and $\mathbf{I}^*$ using $O_i^* - I_i^* \sim \mathcal{G}$

and accept/reject the new augmented data with probability $\max(1, \frac{p(\mathbf{d},\mathbf{O}^*,\mathbf{I}^*|\theta)}{p(\mathbf{d},\mathbf{O},\mathbf{I}|\theta)})$ where $(\mathbf{O}, \mathbf{I})$ represents the previous augmented data state; in practice, 10% of augmented data are changed as a block at each iteration yielding acceptance rates of 10-50%.

2. Propose a new value $R_0^*$ using $R_0^* \sim \mathcal{N}(R_0, \sigma_1)$ where $\mathcal{N}$ is the normal distribution, $R_0$ represents the previous parameter state, and $\sigma_1$ is the fixed standard deviation of the proposal; accept/reject $R_0^*$ as the new current state with probability $\max(1, \frac{p(R_0^*,\lambda_z|x)}{p(R_0,\lambda_z|x)})$. $\sigma_1$ was manually tailored to 0.4 to ensure acceptance rates of 10-50%.
3. Propose a new value $\lambda_z^*$ using $\lambda_z^* \sim \mathcal{N}(\lambda_z, \sigma_2)$ where $\lambda_z$ represents the previous parameter state, and $\sigma_2$ is the fixed standard deviation of the proposal; accept/reject $\lambda_z^*$ as the new current state with probability $\max(1, \frac{p(R_0,\lambda_z^*|x)}{p(R_0,\lambda_z|x)})$. $\sigma_2$ was manually tailored to 0.1 to ensure acceptance rates of 10-50%.
4. Go back to step 1 until desired number of iterations reached

FIGURES

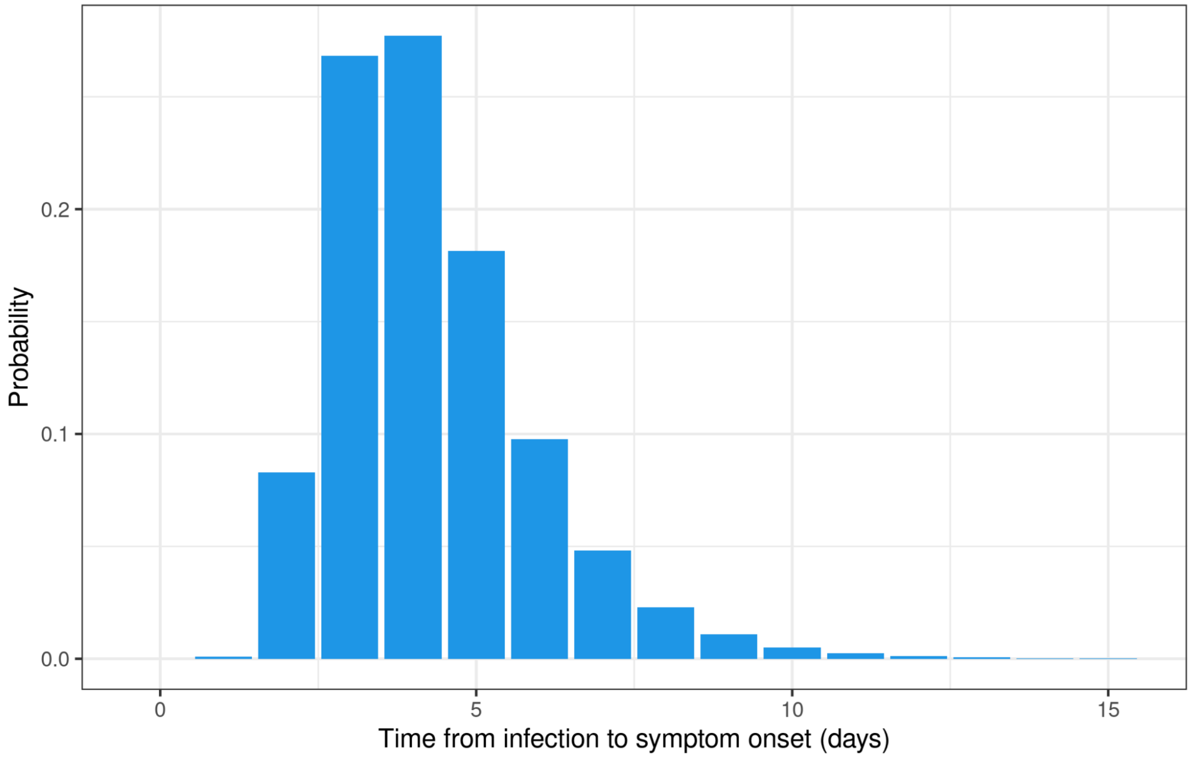


**Figure S1. Incubation time distribution of bubonic plague.** The incubation time distribution was characterized as a discretized log-normal distribution with mean 3.5 days and standard deviation (SD) 1.5 days.

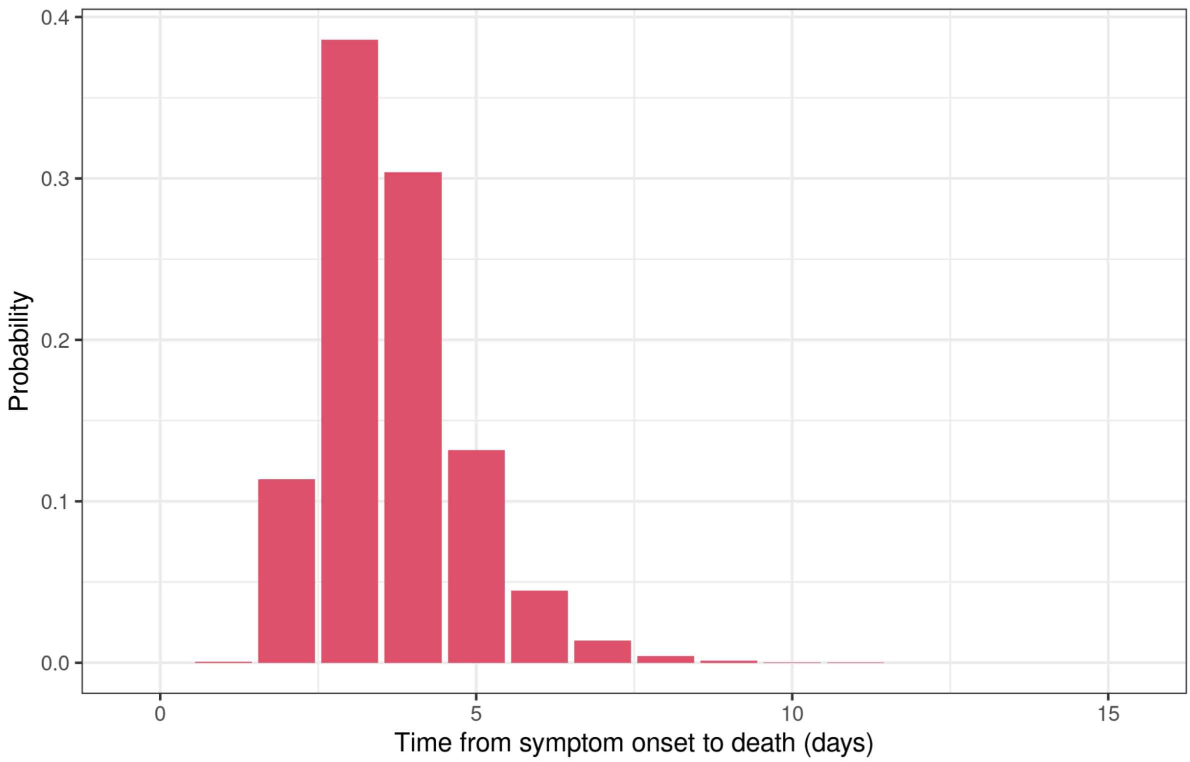


**Figure S2. Duration of illness distribution of bubonic plague.** The incubation time distribution was characterized as a discretized log-normal distribution with mean 3 days and standard deviation (SD) 1.4 days.

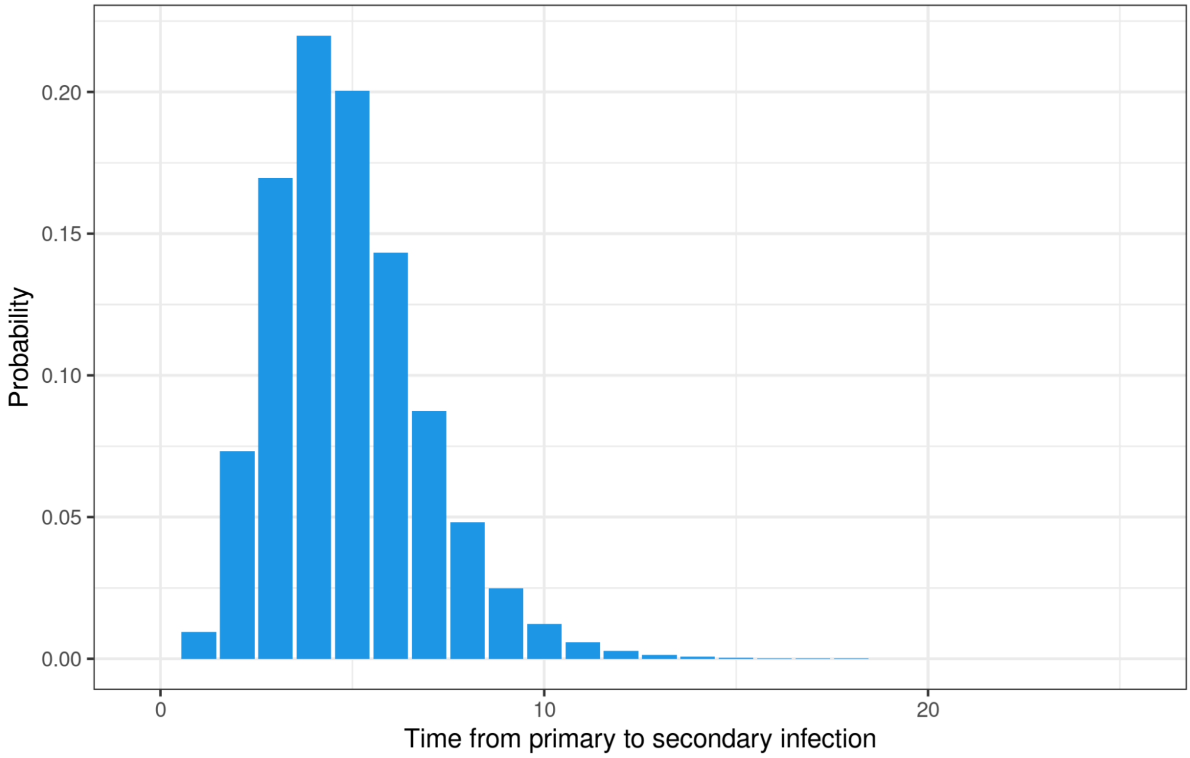


**Figure S3. Generation time distribution of bubonic plague.** This distribution was generated following the procedure described in Text S1, and had mean 4.89 and standard deviation 1.98.

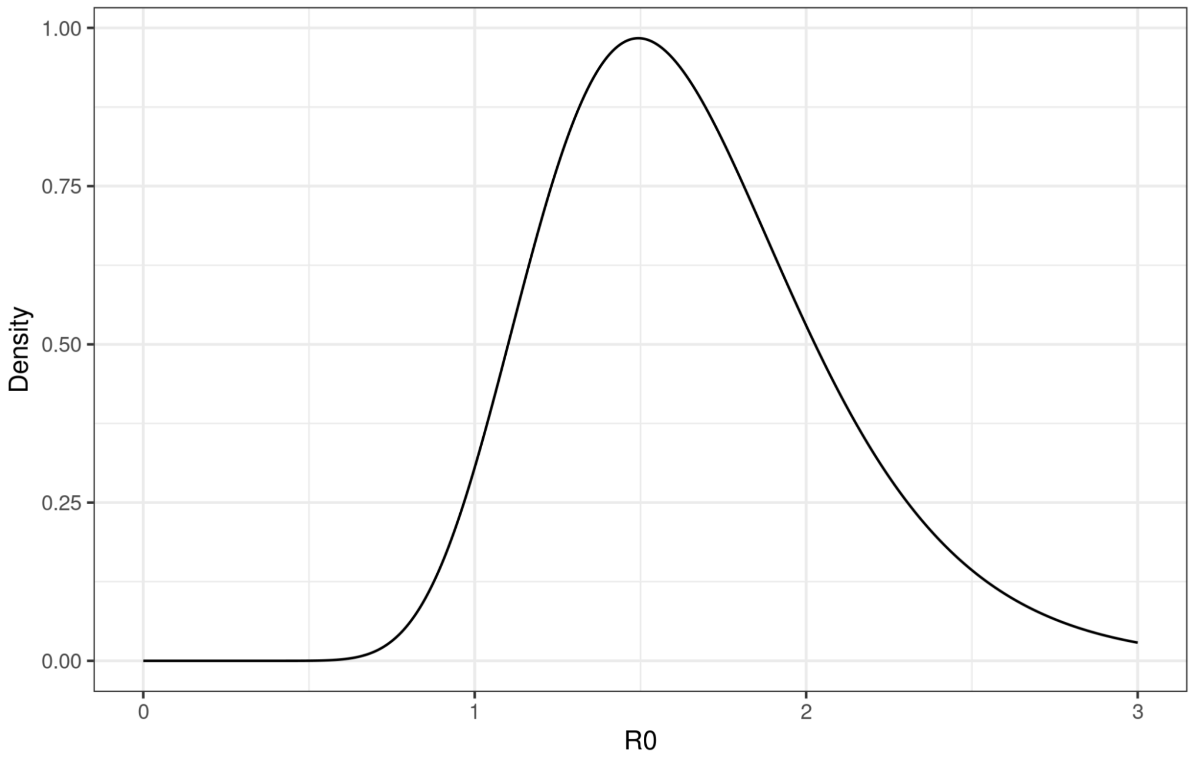


**Figure S4. Prior distribution of the basic reproduction number of bubonic plague, $R_0$.**
This distribution was parameterized as a lognormal with mean 1.6 and SD 1.3, following [1].

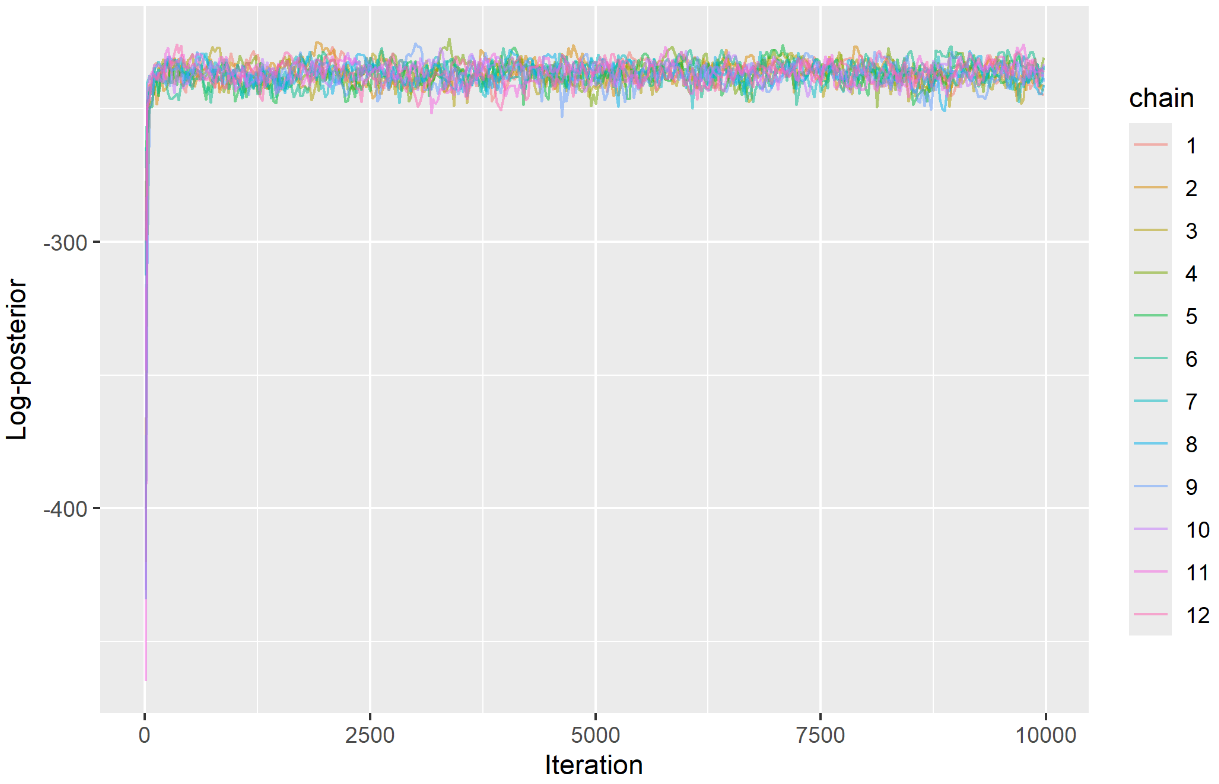


**Figure S5. Trace of log-posterior traces of the transmission model.** Lines represent the log-posterior values of samples derived by the Metropolis algorithm for 12 independent chains (different colors) run in parallel for 10,000 iterations each, after a thinning of 1/25.
